# Tumor Budding Is a Promising Indicator for Elective Neck Dissection in Node-Negative Early-Stage Oral Squamous Cell Carcinoma: A Retrospective Cohort Study

**DOI:** 10.1101/2025.09.16.25334216

**Authors:** Zhuqin Xiang, Bokai Yun, Wenjin Wang, Gan Xiong, Chen Yi, Jinqi Zhang, Yaqi Huang, Linlin Ren, Nan Xie, Zehang Zhuang, Jinsong Hou, Cheng Wang

## Abstract

**Background:** Currently, it is still under debate whether patients with early-stage oral cancers should be treated with elective neck dissection at the time of the primary surgery or with therapeutic neck dissection after cervical nodal relapse. Thus, it is urgent to develop a novel biomarker to stratify the patients with high risk of occult cervical node metastasis who should be treated with elective neck dissection.

**Methods:** A total of 231 patients diagnosed with early-stage oral squamous cell carcinoma were retrospectively retrieved in this cohort study. Tumor budding was evaluated as International Tumor Budding Consensus Conference recommended. Overall and disease-free survivals were evaluated by the Kaplan-Meier method. Cox proportional hazards regression models were used to assess their prognostic value. Subgroup analysis was conducted, and both multiplicative interaction effects and additive interaction effects were assessed.

**Results:** As expected, high tumor budding was associated with poorer prognosis, while elective neck dissection was linked to a better prognosis. After further subgrouping, we were surprised to find that elective neck dissection could significantly improve both overall survival and disease-free survival in patients with high tumor budding, and additive interaction was observed. However, there was no significant difference in overall survival for patients with low tumor budding.

**Conclusion:** Early-stage clinical node-negative oral cancers patients with high tumor budding should be treated with elective neck dissection for better prognosis. The ‘Watch and wait’ and therapeutic neck dissection strategies are still recommended for early-stage oral cancers patients with low tumor budding.

## Introduction

Oral squamous cell carcinoma (OSCC), which accounts for nearly 50% of all head and neck cancers, resulted in over 389,485 new cases and caused 188,230 deaths globally in 2022 alone[1]. One of its characteristic features is a high risk of cervical lymph node metastasis, which is a major source of disease burden and mortality[2]. The conventional OSCC management includes the primary tumor resection and neck dissection. A consensus has been reached that neck dissection should be performed when cervical lymph node metastasis is clinically detected[3]. However, controversy still exists regarding the treatment of patients with early-stage, clinically node-negative OSCC. Currently, such patients usually undergo elective neck dissection (END) while receiving primary tumor resection, or follow the “watch and wait” strategy, and receive therapeutic neck dissection after nodal recurrence is detected.

Over the past decade, increasing evidence has supported the notion that END improves both overall survival and disease-free survival in patients with node-negative early-stage oral cancer[4, 5]. Moreover, a cost-effectiveness analysis showed that performing END for early-stage OSCC can reduce treatment costs over a lifetime by reducing the use of salvage therapy[6]. However, the reported mean incidence of subclinical occult metastasis in such node-negative early-stage OSCC patients was 25.9% at the initial visit[7]. Blindly performing neck dissection may lead to 60-80% of patients undergoing overtreatment[8], which may can cause dysfunction in the head, neck, and shoulders and damage local immune function[6]. Thus, there is currently a lack of an adjunctive diagnostic method to stratify high-risk patients who require END and assist surgeons in making precise treatment decisions.

Tumor budding (TB) is typically defined as isolated single cancer cells or a small tumor cell cluster of up to four cancer cells, located at the tumor invasive front[9]. It is a pathological feature in which tumor buds detach from the main tumor mass and infiltrate into the adjacent stroma, indicating that the tumor cells possess strong invasiveness and metastatic potential[9, 10]. Biologically, it is considered a part of the tumor microenvironment (TME) and is a morphologic characteristic of epithelial-mesenchymal transition (EMT) or partial EMT (p-EMT) [10, 11].

Currently, tumor budding has been recognized as an emerging prognostic biomarker that can predict disease progression and survival in several solid cancers, especially in colon cancer[12–14]. The value of TB in clinical decision-making has been confirmed in colon cancer. It is an adverse prognostic factor in determining whether radical surgery is needed for pT1 colorectal cancer patients and whether adjuvant chemotherapy is needed for stage II colorectal cancer patients[15, 16]. Compared with colorectal cancer, there is relatively less research on the tumor budding in oral cancer despite of its prognostic value validated in our previous studies [17–19] and other studies[12, 20]. Moreover, the role of tumor budding in treatment planning for oral cancer is still not evaluated.

In this study, we analyzed the relationship between TB, END, and clinical outcomes in patients with early-stage, clinically node-negative OSCC. We explored the prognostic value of TB and assessed whether it could serve as a reference indicator for surgical decision-making, thus providing additional guidance for clinical treatment strategies.

## Materials and Methods

### Study Cohort

Overall, this retrospective study included 231 patients with early-stage OSCC from the Hospital of Stomatology, Sun Yat-sen University, covering the period from January 2010 to December 2017. The inclusion criteria included: (1) Patients who had not received any previous treatment and had no history of head and neck cancer; (2) Patients with available diagnostic slides and histopathologically confirmed as T1 or T2 OSCC; (3) Patients who had no clinical evidence of nodal involvement in the neck; (4) Patients with complete medical records and who had access to follow-up. Exclusion criteria included: (1) Patients with other malignant tumors, such as esophageal cancer; (2) Patients lacking pathological slides with tumor invasive front or exhibiting poor staining of slides. Moreover, due to all patients were staged with American Joint Committee on Cancer (AJCC) seventh edition at the time of their treatment, which was outdated. We restaged according to the AJCC eighth edition staging system, and those who staged as higher T classification were excluded.

This retrospective cohort study received approval from the ethical committee of Sun Yat-Sen University and Guanghua School of Stomatology. Due to the retrospective and deidentified data utilize in this study, the informed consent was waived by committee. This cohort study has been reported in line with the Strengthening the Reporting of Observational Studies in Epidemiology (STROBE) reporting guidelines.

### Follow-up and clinical outcome

The clinical data, including survival and recurrence status, were acquired from patient follow-up medical records and telephone follow-up. The primary end point was overall survival (OS), which was defined as the time from surgical resection to death from any cause, with censoring at last follow-up. The secondary end point was disease-free survival (DFS), which was defined as the time from surgical resection to relapse at any site (local, regional or distal) or death from any cause.

### Histopathological assessment

This study used the hematoxylin and eosin (H&E) stained slides that archived in the Department of Oral Pathology, Hospital of Stomatology for pathological analysis. The integrity, color of the tissue, and existence of the tumor invasive front were checked. All slides were scanned and assessed using Aperio ScanScope AT 2, eslide Manager and ImageScope software (Leica Biosystems).

Two trained authors who were blinded to the clinical data reviewed all the pathological specimens independently. The pathological differentiation was classified according to the WHO classification. The depth of invasion was measured from the surface of the mucosa to the deepest point of the tumor.

TB was assessed according to the recommendations of the International Tumor Budding Consensus Conference (ITBCC) [15]. Briefly, the invasive front of all slides was scanned at a low power objective initially to find the hotspot with the greatest density of tumor budding. Then, the number of tumor budding were counted in an area of 0.785 mm^2^ on the hotspots at 20× magnification, and grading the TB grade as Bd1 (0-4 buds), Bd2 (5-9 buds) and Bd3 (buds≥10). Referring to other relevant studies[20], in this study, Bd2 and Bd3 was considered high grade, while Bd1 was considered low grade. Cases for which a determination of TB grade was challenging, two authors reviewed together and achieved consensus. Kappa statistics was performed to assess the interobserver variability and achieved great agreement (K = 0.814).

### Statistical analyses

Kaplan-Meier method was used to estimate the OS and DFS after primary tumor resection and tested by means of log-rank tests. Cox proportional hazards regression model was used to assess the effect of multiple variables and report hazard ratios (HRs) with 95% confidence intervals (CIs). Additionally, we conducted subgroup analyses based on TB and other clinicopathological features known to have an effect on survival, which was performed using a Cox proportional hazards mode. Interaction tests, including both multiplicative and additive interactions, were also performed. The “mover” method was used to calculate the 95% confidence interval of additive interaction. Additive interactions assessed by ‘interactionR’ package. Preventive or protective exposure factors were recoded to ensure consistency in the direction of the effects, which helped standardize the direction of additive interaction effects analysis.

Statistical analyses were performed using RStudio (version 4.2.3; RStudio Inc., Boston, MA). The following R packages were used: ‘survival’ and ‘plyr’ for cox regression analysis, ‘survminer’ for survival curves generation, ‘interactionR’ for interaction tests, ‘ggplot2’ and ‘forestploter’ for data visualization. All statistical tests with P-values <0.05 (2 tailed) were considered significant. Data were analyzed from July to October 2024.

## Results

### Clinicopathologic characteristics of the patients

As shown in **Table 1**, a total of 231 patients were enrolled in this study. Among them, 138 (59.7%) were male, and 93 (40.3%) were female. The median age of patients at diagnosis was 54 years (range, 19 to 87 years). Tongue was the most common site of tumors, accounting for 147 cases (63.6%), followed by the buccal mucosa with 34 cases (14.7%). There were 89 (38.5%) patients classified as pT1, while 142 (61.5%) patients classified as pT2. The number of patients with pathological differentiation of well and moderately/poorly were 119 (51.5%) and 112 (48.5%), respectively. Tumor buds can be readily identified based on standard H&E staining as shown in **Figure 1**. The number of tumor buds in the slides ranged from 0 to 40 (median, 5). As mentioned earlier, on the basis of the budding grade, 103 (44.6%) and 128 (55.4%) patients were classified as low budding group and high budding group, respectively. In this cohort, 156 patients (67.5%) underwent END concurrent with primary tumor resection, while 75 (32.5%) only underwent primary tumor resection and subsequent observation (OBS). Of these 156 patients, 15 (9.6%) cases were positive lymph node status.

**Figure 1:**
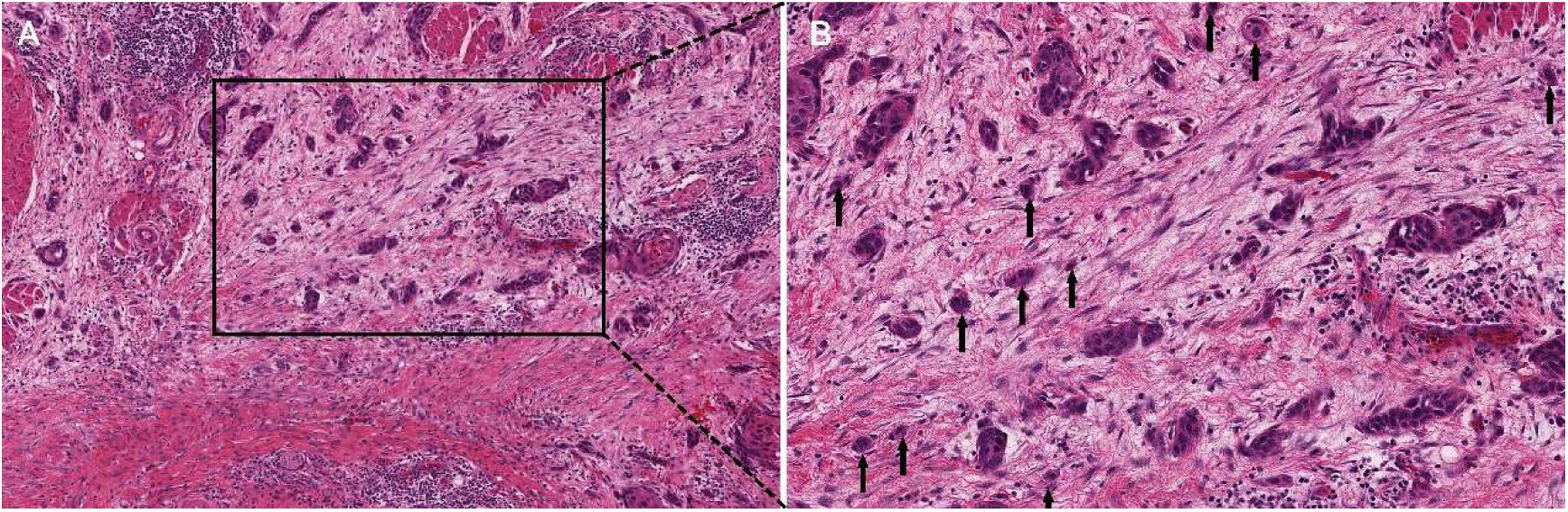
Tumor Budding at The Invasive Front in Oral Squamous Cell Carcinoma. Arrowheads indicate tumor buds. (Hematoxylin–eosin stain; original magnifications: A,×20; B,×40.)

**Table 1.**
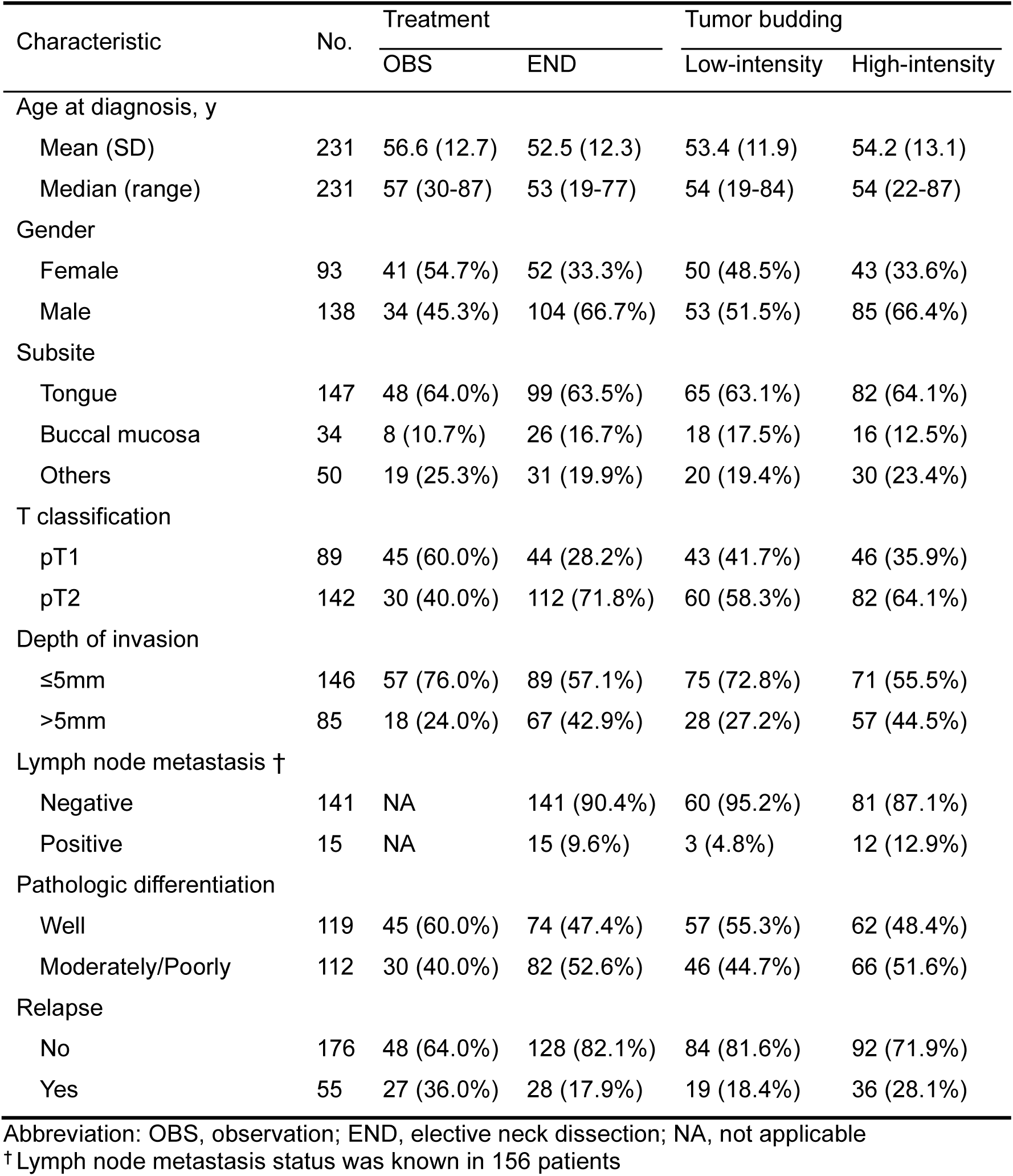
Clinicopathologic Characteristics of the Study Cohort.

The follow-up time for the cohort ranged from 6 to 123 months, with a median follow-up time of 75 months. At the time of last follow-up, 163 (70.6%) patients were alive with no evidence of relapse, 55 (23.8%) patients had recurred, 33 (14.3%) patients had died of any causes. Among the recurrent patients, 26 (47.3%) cases had local recurrence, 18 (32.7%) cases had regional lymph nodes metastasis, 2 (3.6%) cases had distant metastasis, 6 (10.9%) cases had both local recurrence and regional lymph nodes metastasis, and 3 (5.5%) cases had both regional lymph nodes metastasis and distant metastasis. The median of local recurrence, regional lymph nodes metastasis and distant metastasis were 26 months, 7 months and 12 months, respectively.

### Survival analysis

On the basis of the TB grade, the 5-year OS rate of the high budding group was 80.4% (95% CI, 73.8-87.6), which was significantly lower than 93% (95% CI, 88.2-98.1) of the low budding group (p = 0.0059; **Figure 2A**). The corresponding 5-year DFS rates showed a similar association, with a rate of 66.2% (95% CI, 58.4-79.4) in the high budding group, significantly lower than 80.6% (95% CI, 73.3-88.6) in the low budding group (p = 0.019; **Figure 2B**). Similarly, on the basis of treatment, the 5-year OS rate of the END group was 90.3% (95% CI, 85.5-95.1), significantly higher than 76.8% (95% CI, 67.7-87.1) of the OBS group (p = 0.0058; **Figure 2C**). The 5-year DFS rate of the END group was 81.3% (95% CI, 75.4-87.7), which was significantly higher than 54.6% (95% CI, 44.4-67.2) of the OBS group (p < 0.0001; **Figure 2D**).

**Figure 2:**
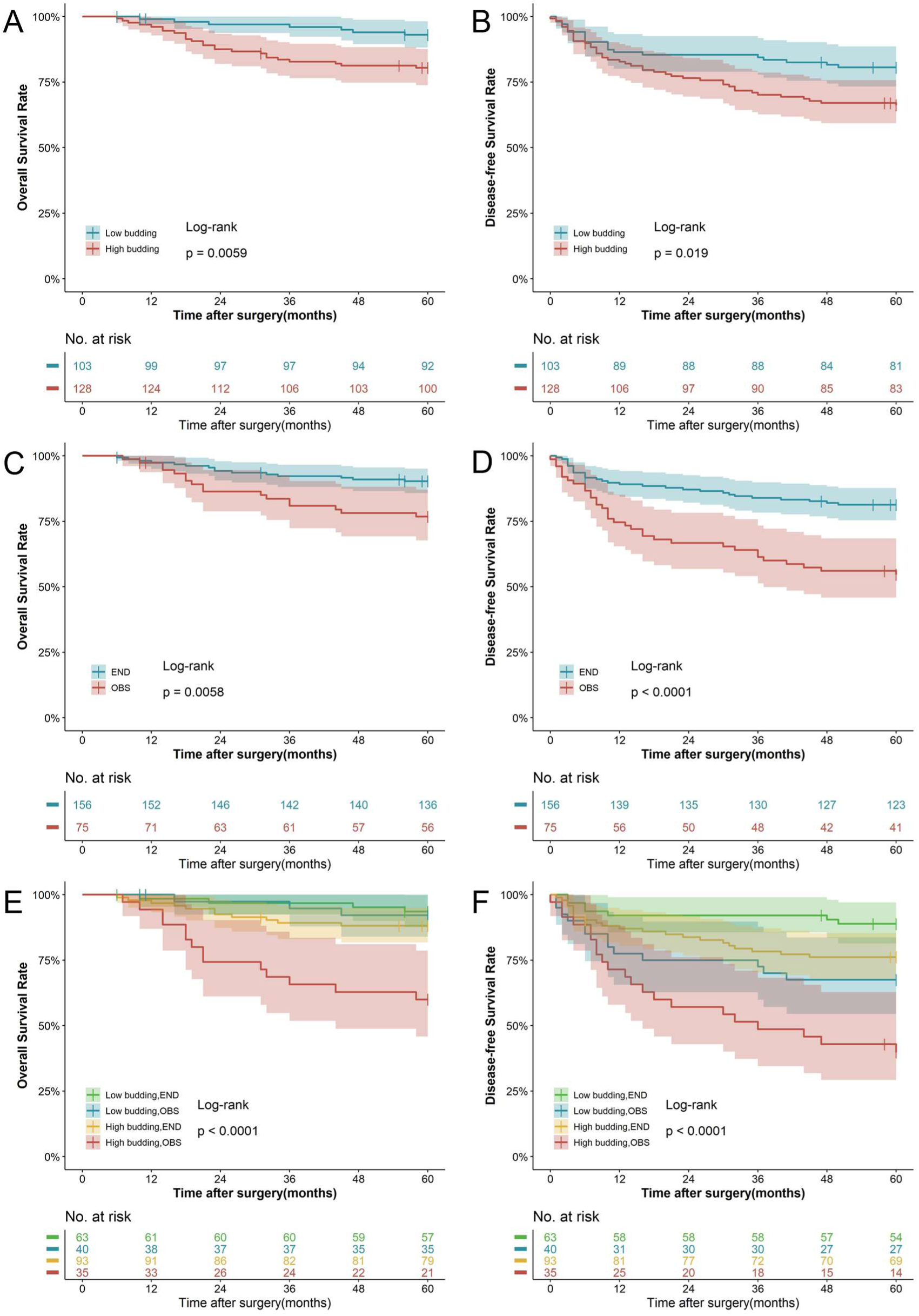
Kaplan-Meier Estimates of Overall Survival and Disease-Free Survival Kaplan–Meier estimates of OS and DFS stratified by tumor budding grade (A, B); Kaplan–Meier estimates of OS and DFS stratified by treatment (C, D); Kaplan–Meier estimates of OS and DFS stratified by tumor budding grade combined with treatment (E, F). Colored areas represent a 95% confidence interval. Abbreviations: OS, overall survival; DFS, disease-free survival; END, elective neck dissection; OBS, observation.

Furthermore, both univariate and multivariate Cox regression analyses were performed on established clinicopathological parameters. As shown in **Table 2**, high grade of TB (HR = 3.67, 95% CI: 1.66-8.09, p = 0.001) indicated a poor prognosis, while END (HR = 0.28, 95% CI: 0.14-0.54, p < 0.001) indicated a favorable prognosis in multivariate analysis for OS. Similarly, as shown in **Table 3**, high TB grade predicted a poor prognosis (HR = 2.19, 95% CI: 1.30-3.70, p = 0.003), while END indicated a favorable prognosis in multivariate analysis for DFS (HR = 0.35, 95% CI: 0.21-0.56, p < 0.001).

**Table 2.** Cox Regression Model for Overall Survival.

| Characteristic | Univariate analysis |  |  | Multivariate analysis |  |  |
| --- | --- | --- | --- | --- | --- | --- |
|  | HR | 95% CI | P value | HR | 95% CI | p |
| Age (years) |  |  |  |  |  |  |
| <60 | 1 |  |  | 1 |  |  |
| ≥60 | 2.30 | 1.21-4.38 | 0.011 | 2.04 | 1.07-3.89 | 0.032 |
| Gender |  |  |  |  |  |  |
| Female | 1 |  |  |  |  |  |
| Male | 0.98 | 0.51-1.89 | 0.950 |  |  |  |
| Subsite |  |  |  |  |  |  |
| Others | 1 |  |  |  |  |  |
| Tongue | 0.98 | 0.50-1.91 | 0.954 |  |  |  |
| T classification |  |  |  |  |  |  |
| pT1 | 1 |  |  |  |  |  |
| pT2 | 1.19 | 0.61-2.34 | 0.615 |  |  |  |
| Depth of invasion |  |  |  |  |  |  |
| ≤5mm | 1 |  |  |  |  |  |
| >5mm | 1.05 | 0.53-2.06 | 0.888 |  |  |  |
| Pathologic differentiation |  |  |  |  |  |  |
| Well | 1 |  |  |  |  |  |
| Moderately/Poorly | 1.11 | 0.58-2.13 | 0.746 |  |  |  |
| END |  |  |  |  |  |  |
| No | 1 |  |  | 1 |  |  |
| Yes | 0.33 | 0.17-0.63 | 0.001 | 0.28 | 0.14-0.54 | <0.001 |
| TB |  |  |  |  |  |  |
| Low budding | 1 |  |  | 1 |  |  |
| High budding | 3.16 | 1.44-6.92 | 0.004 | 3.67 | 1.66-8.09 | 0.001 |

Abbreviation: HR, hazard ratio; CI, confidence interval; END, elective neck dissection; TB, tumor budding

**Table 3.** Cox Regression Model for Disease-Free Survival.

| Characteristic | Univariate analysis |  |  | Multivariate analysis |  |  |
| --- | --- | --- | --- | --- | --- | --- |
|  | HR | 95% CI | *p* | HR | 95% CI | *p* |
| Age(years) |  |  |  |  |  |  |
| <60 | 1 |  |  | 1 |  |  |
| ≥60 | 1.74 | 1.07-2.81 | 0.024 | 1.56 | 0.96-2.53 | 0.074 |
| Gender |  |  |  |  |  |  |
| Female | 1 |  |  |  |  |  |
| Male | 0.78 | 0.48-1.25 | 0.296 |  |  |  |
| Subsite |  |  |  |  |  |  |
| Others | 1 |  |  |  |  |  |
| Tongue | 0.75 | 0.47-1.22 | 0.250 |  |  |  |
| T classification |  |  |  |  |  |  |
| pT1 | 1 |  |  |  |  |  |
| pT2 | 1.17 | 0.62-1.66 | 0.949 |  |  |  |
| Depth of invasion |  |  |  |  |  |  |
| ≤5mm | 1 |  |  |  |  |  |
| >5mm | 1.20 | 0.62-1.66 | 0.470 |  |  |  |
| Pathologic differentiation |  |  |  |  |  |  |
| Well | 1 |  |  |  |  |  |
| Moderately/Poorly | 1.17 | 0.73-1.89 | 0.513 |  |  |  |
| END |  |  |  |  |  |  |
| No | 1 |  |  | 1 |  |  |
| Yes | 0.38 | 0.23-0.61 | <0.001 | 0.35 | 0.21-0.56 | <0.001 |
| TB |  |  |  |  |  |  |
| Low budding | 1 |  |  | 1 |  |  |
| High budding | 1.97 | 1.18-3.30 | 0.010 | 2.19 | 1.30-3.70 | 0.003 |

Abbreviation: HR, hazard ratio; CI, confidence interval; END, elective neck dissection; TB, tumor budding

Since both TB and END were independent factors in the multivariate analysis, we subdivided the cohort into 4 groups on the basis of the tumor budding grade and treatment, the 5-year OS curve and DFS curve both show significant difference (p < 0.0001, p < 0.0001, respectively; **Figure 2E, 2F**), the patients with high budding grade and only receive primary tumor resection had the worse prognosis.

In patients with high budding grade, the rate of 5-year OS and DFS for the END group were 88.1% (95% CI, 81.8-95.0) and 76.2% (95% CI, 68.0-85.4), respectively, both significantly higher than those of the OBS group, which were 60% (95% CI, 45.8-78.6) and 39.8% (95% CI, 26.4-59.9), respectively. (p = 0.00024, p < 0.0001, respectively; **Figure S1A and S2A** in Supplement 1). In patients with low budding grade, the 5-year DFS rate in the END group was 88.8% (95% CI, 81.4-97.0), significantly higher than 67.5% (95% CI, 54.4-83.7) in the OBS group, while the 5-year OS rate were 93.5% (95% CI, 87.6-99.9) and 92.1% (95% CI, 83.9-100.0), respectively, shows no significant difference. (p < 0.0001, p = 0.79, respectively; **Figure S2B and S1B** in Supplement 1).

### Subgroup analysis

Furthermore, we conducted a subgroup analysis to explore the differences in the effects of END across various subgroups of the cohort. As shown in **Figure 3**, stratified by clinicopathologic factors, the overall survival benefits of END were seen across prespecified subgroups. However, in each subgroup, the test of multiplicative interaction was not significant. Subsequently, following recent recommendations[21], we further assess the additive interactions, which showed that in the subgroup stratified by TB, there was a significant additive interaction between TB and END (RERI = 7.12 [95% CI, 1.8-24.9], AP = 0.72 [95% CI, 0.27-0.94], SI = 4.92 [95% CI, 1.02-23.78]). For other stratified factors, there was no significant additive interaction effect between them and END.

**Figure 3:**
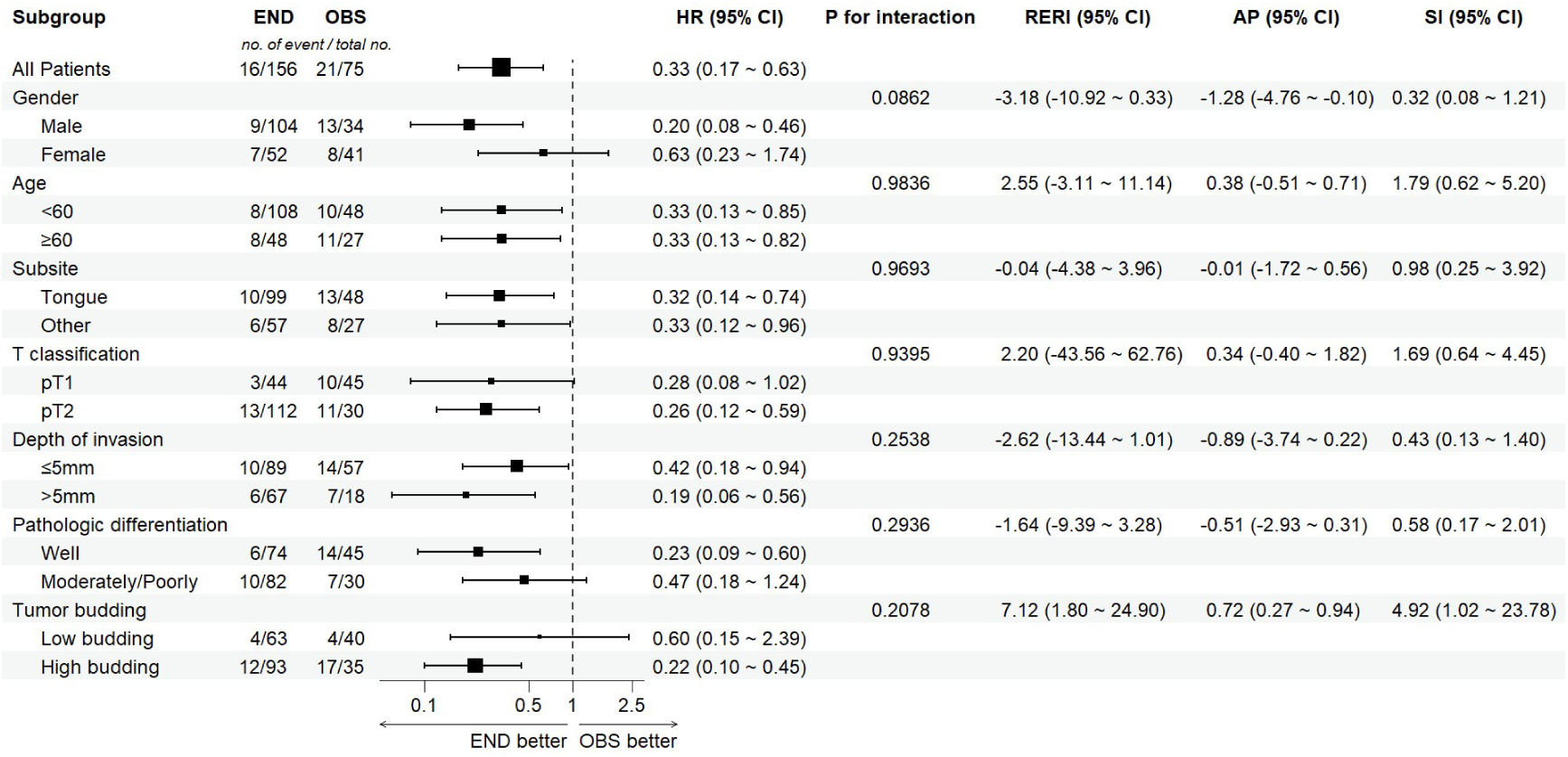
Subgroup Analyses of Overall Survival. The size of the squares corresponds to the number of patients with an event. A HR of <1.00 indicates a lower risk of death with elective neck dissection than with observation. Additive interaction was statistically significant when the RERI 95% CI and AP 95% CI did not include 0, and the SI 95% CI did not include 1. If both RERI and AP were greater than 0, and SI was greater than 1, there was a synergistic effect between the two factors. If both RERI and AP were less than 0, and SI was less than 1, there was an antagonistic effect between the two factors. Abbreviations: END, elective neck dissection; OBS, observation; CI, confidence interval; HR, hazard ratio; RERI, relative excess risk due to interaction; AP, attributable proportion due to interaction; SI, the synergy index.

## Discussion

Currently, the treatment of OSCC is based on the clinical TNM staging system, and predicting the prognosis of early-stage OSCC patients is one of the biggest challenges. One of the reasons is the high incidence of occult metastases and recurrence in the neck, which cannot be confirmed by routine imaging examinations[7]. Some prospective randomized controlled trial and meta-analyses suggest that patients who received END at the time of primary tumor resection had better overall survival, a lower likelihood of nodal recurrence[4, 5, 7, 22]. As expected, our research also yielded similar results, showing that in early-stage, clinically node-negative OSCC patients, the overall survival rate and disease-free survival rate of the END group increasing by 13.5 percentage points and 26.7 percentage points, respectively, compared to the observation group. However, some of other studies did not support the above viewpoint[23, 24]. The necessity of END for early-stage OSCC still lacks conclusive evidence, and the “watch and wait” strategy is still recommended in the US National Comprehensive Cancer Network (NCCN) 2024 guidelines and other widely followed guidelines[3]. Therefore, using more sensitive and specific prognostic biomarkers to stratify the risk of OSCC patients can be of clinical significance.

Tumor budding is commonly observed in several solid cancer, and increasing evidence suggests that TB is an emerging prognostic biomarker, with a negative impact on disease prognosis and survival[12–14]. Consistent with the above finding, in our study cohort, the overall survival rate and disease-free survival rate of patients with high grade tumor budding decreased by 12.6 percentage points and 14.4 percentage points as compared to the patients with low grade tumor budding, respectively. The adverse prognostic impact of tumor budding is considered attributed to its association with the EMT and p-EMT, which play a critical role in the metastatic cascade of cancer[10]. Multiple studies have shown that the expression of E-cadherin, a cell–cell adhesion protein and crucial regulator of EMT, is reduced at the invasive tumor front and especially within tumor buds[25–27]. In OSCC, tumor buds have been shown to overexpress SNAIL and TWIST1 as well as ZEB1, the EMT-associated transcription factors[26, 28]. Our previous researches also indicated that tumor budding displayed EMT or p-EMT phenotype and gene signatures based immunohistochemistry, laser capture microdissection and scRNA-seq analysis[11, 17].

Currently, accumulated evidence suggests that more aggressive treatment strategies may be warranted for patients with high grade TB. It is an unfavorable prognostic factor that helps determine whether patients with pT1 colorectal cancer require radical surgery and whether patients with stage II colorectal cancer should undergo adjuvant chemotherapy[15, 16]. In this study, the prognostic value of TB and END was shown to be independent in early-stage clinically node-negative OSCC. Patients with high-grade TB who did not undergo END had the worst prognosis in both OS and DFS. Interestingly, for patients with low-grade TB, there was no significant difference in OS between those who received END and those who did not. In subsequent subgroup analysis, TB and END did not exhibit a significant multiplicative interaction effect. However, following the recommendation of Mansournia et al.[21], additive interaction on risks is more relevant for both clinical decisions and public health and therefore should be assessed. As a result, we also conducted an additive interaction effect analysis, which revealed a significant additive interaction between END and TB. This suggests that TB grade has the potential to serve as a criterion for END in patients with early-stage clinically node-negative OSCC. Although samples in this study were all postoperative pathological slides, it is feasible to assess tumor budding through preoperative biopsy or intraoperative frozen section[29]. Currently limited research indicates that intraoperative frozen sections are relatively difficult to identify individual cells or cell nests due to the presence of artifacts, resulting in poor accuracy in assessing budding grade[30]. Anyway, for such patients, if preoperative pathological biopsy shows a high density of tumor budding at the tumor invasive front, END may provide significant clinical benefits in terms of both OS and DFS.

There are some limitations in our study. This study is a single center, retrospective study, due to surgeon bias and patient autonomy choice, there are inherent biases present in such studies, and the results may not be fully representative of other populations or health care settings. For this reason, our findings require validation from other centers, ideally in prospective designed studies. The patients in this study cohort underwent different preoperative evaluations. Additionally, since clinically negative lymph nodes are a relative concept, the accuracy of preoperative imaging for detecting occult lymph node metastasis has improved with the further development of deep learning and radiology[31]. This could result in discrepancies between the recorded clinical staging and the actual condition.

## Conclusions

In conclusion, the results of our study suggest that early-stage oral cancers patients (pT_1/2_cN_0_M_0_) with high tumor budding may achieve benefits in overall survival and disease-free survival if performing elective neck dissection at the same time of primary tumor resection. The “Watch and wait” and therapeutic neck dissection strategies are still recommended for early-stage oral cancers patients with low tumor budding.

## Data Availability

All data produced in the present study are available upon reasonable request to the authors

## Acknowledgments

This work was supported by the National Natural Science Foundation of China (82473072, 82073265).

## Data statement

The data that support the findings of this study are available from the corresponding author upon reasonable request.

## References

[1] Bray F, Laversanne M, Sung H, Ferlay J, Siegel RL, Soerjomataram I, et al. Global cancer statistics 2022: GLOBOCAN estimates of incidence and mortality worldwide for 36 cancers in 185 countries. CA Cancer J Clin. 2024;74:229–63.

[2] Woolgar JA, Rogers SN, Lowe D, Brown JS, Vaughan ED. Cervical lymph node metastasis in oral cancer: the importance of even microscopic extracapsular spread. Oral Oncol. 2003;39:130–7.

[3] National Comprehensive Cancer Network. Head and Neck Cancers (Version 5.2024).

[4] D’Cruz AK, Vaish R, Kapre N, Dandekar M, Gupta S, Hawaldar R, et al. Elective versus Therapeutic Neck Dissection in Node-Negative Oral Cancer. N Engl J Med. 2015;373:521–9.

[5] Hutchison IL, Ridout F, Cheung SMY, Shah N, Hardee P, Surwald C, et al. Nationwide randomised trial evaluating elective neck dissection for early stage oral cancer (SEND study) with meta-analysis and concurrent real-world cohort. Br J Cancer. 2019;121:827–36.

[6] Acevedo JR, Fero KE, Wilson B, Sacco AG, Mell LK, Coffey CS, et al. Cost-Effectiveness Analysis of Elective Neck Dissection in Patients With Clinically Node-Negative Oral Cavity Cancer. J Clin Oncol. 2016;34:3886–91.

[7] Worthington HV, Bulsara VM, Glenny AM, Clarkson JE, Conway DI, Macluskey M. Interventions for the treatment of oral cavity and oropharyngeal cancers: surgical treatment. Cochrane Database Syst Rev. 2023;8:CD006205.

[8] de Bree R, Takes RP, Shah JP, Hamoir M, Kowalski LP, Robbins KT, et al. Elective neck dissection in oral squamous cell carcinoma: Past, present and future. Oral Oncol. 2019;90:87–93.

[9] Lugli A, Zlobec I, Berger MD, Kirsch R, Nagtegaal ID. Tumour budding in solid cancers. Nat Rev Clin Oncol. 2021;18:101–15.

[10] Zlobec I, Lugli A. Tumour budding in colorectal cancer: molecular rationale for clinical translation. Nat Rev Cancer. 2018;18:203–4.

[11] Wang W, Yun B, Hoyle RG, Ma Z, Zaman SU, Xiong G, et al. CYTOR Facilitates Formation of FOSL1 Phase Separation and Super Enhancers to Drive Metastasis of Tumor Budding Cells in Head and Neck Squamous Cell Carcinoma. Adv Sci (Weinh). 2024;11:e2305002.

[12] Silva F, Caponio VCA, Perez-Sayans M, Padin-Iruegas ME, Mascitti M, Chamorro-Petronacci CM, et al. Tumor budding is a prognostic factor in head and neck squamous cell carcinoma: A comprehensive meta-analysis and trial sequential analysis. Crit Rev Oncol Hematol. 2024;193:104202.

[13] Huang T, Bao H, Meng YH, Zhu JL, Chu XD, Chu XL, et al. Tumour budding is a novel marker in breast cancer: the clinical application and future prospects. Ann Med. 2022;54:1303–12.

[14] Ueno H, Ishiguro M, Nakatani E, Ishikawa T, Uetake H, Matsuda C, et al. Prospective Multicenter Study on the Prognostic and Predictive Impact of Tumor Budding in Stage II Colon Cancer: Results From the SACURA Trial. J Clin Oncol. 2019;37:1886–94.

[15] Lugli A, Kirsch R, Ajioka Y, Bosman F, Cathomas G, Dawson H, et al. Recommendations for reporting tumor budding in colorectal cancer based on the International Tumor Budding Consensus Conference (ITBCC) 2016. Mod Pathol. 2017;30:1299–311.

[16] Baxter NN, Kennedy EB, Bergsland E, Berlin J, George TJ, Gill S, et al. Adjuvant Therapy for Stage II Colon Cancer: ASCO Guideline Update. J Clin Oncol. 2022;40:892–910.

[17] Wang C, Huang H, Huang Z, Wang A, Chen X, Huang L, et al. Tumor budding correlates with poor prognosis and epithelial-mesenchymal transition in tongue squamous cell carcinoma. J Oral Pathol Med. 2011;40:545–51.

[18] Xie N, Yu P, Liu H, Liu X, Hou J, Chen X, et al. Validation of the International Tumor Budding Consensus Conference (2016) recommendations in oral tongue squamous cell carcinoma. J Oral Pathol Med. 2019;48:451–8.

[19] Zhu Y, Liu H, Xie N, Liu X, Huang H, Wang C, et al. Impact of tumor budding in head and neck squamous cell carcinoma: A meta-analysis. Head Neck. 2019;41:542–50.

[20] Almangush A, Pirinen M, Heikkinen I, Makitie AA, Salo T, Leivo I. Tumour budding in oral squamous cell carcinoma: a meta-analysis. Br J Cancer. 2018;118:577–86.

[21] Mansournia MA, Nazemipour M. Recommendations for accurate reporting in medical research statistics. Lancet. 2024;403:611–2.

[22] Wushou A, Wang M, Yibulayin F, Feng L, Lu MM, Luo Y, et al. Patients With cT1N0M0 Oral Squamous Cell Carcinoma Benefit From Elective Neck Dissection: A SEER-Based Study. J Natl Compr Canc Netw. 2021;19:385–92.

[23] Chien CY, Wang CP, Lee LY, Lee SR, Ng SH, Kang CJ, et al. Indications for elective neck dissection in cT1N0M0 oral cavity cancer according to the AJCC eight edition: A nationwide study. Oral Oncol. 2023;140:106366.

[24] Yuen AP, Ho CM, Chow TL, Tang LC, Cheung WY, Ng RW, et al. Prospective randomized study of selective neck dissection versus observation for N0 neck of early tongue carcinoma. Head Neck. 2009;31:765–72.

[25] Kohler I, Bronsert P, Timme S, Werner M, Brabletz T, Hopt UT, et al. Detailed analysis of epithelial-mesenchymal transition and tumor budding identifies predictors of long-term survival in pancreatic ductal adenocarcinoma. J Gastroenterol Hepatol. 2015;30 Suppl 1:78–84.

[26] Jensen DH, Dabelsteen E, Specht L, Fiehn AM, Therkildsen MH, Jonson L, et al. Molecular profiling of tumour budding implicates TGFbeta-mediated epithelial-mesenchymal transition as a therapeutic target in oral squamous cell carcinoma. J Pathol. 2015;236:505–16.

[27] Nakagawa Y, Ohira M, Kubo N, Yamashita Y, Sakurai K, Toyokawa T, et al. Tumor budding and E-cadherin expression are useful predictors of nodal involvement in T1 esophageal squamous cell carcinoma. Anticancer Res. 2013;33:5023–9.

[28] Hong KO, Oh KY, Shin WJ, Yoon HJ, Lee JI, Hong SD. Tumor budding is associated with poor prognosis of oral squamous cell carcinoma and histologically represents an epithelial-mesenchymal transition process. Hum Pathol. 2018;80:123–9.

[29] Urken ML, Yun J, Saturno MP, Greenberg LA, Chai RL, Sharif K, et al. Frozen Section Analysis in Head and Neck Surgical Pathology: A Narrative Review of the Past, Present, and Future of Intraoperative Pathologic Consultation. Oral Oncol. 2023;143:106445.

[30] Cacchi C, Fischer HJ, Wermker K, Rashad A, Jonigk DD, Holzle F, et al. New Tumor Budding Evaluation in Head and Neck Squamous Cell Carcinomas. Cancers (Basel). 2024;16.

[31] Lan T, Kuang S, Liang P, Ning C, Li Q, Wang L, et al. MRI-based deep learning and radiomics for prediction of occult cervical lymph node metastasis and prognosis in early-stage oral and oropharyngeal squamous cell carcinoma: a diagnostic study. Int J Surg. 2024;110:4648–59.

